# Systematic violations of patients’ rights and safety: Forced medication of a cohort of 30 patients in Alaska

**DOI:** 10.1101/2022.11.22.22282650

**Authors:** Gail Tasch, Peter C Gøtzsche

## Abstract

We assessed the records for 30 consecutive petitions for mental health commitment in which an involuntary medication order was requested from Anchorage, Alaska. In 29 cases, the commitment petition was granted. One patient requested a jury trial and the jury found in her favor. The forced medication order was granted in 27 of the 30 cases. In 26 cases, in violation of previous Supreme Court rulings, the patients’ desires, fears, wishes and experiences were totally ignored even when the patients were afraid that the drugs used for psychosis might kill them or when they had experienced serious harms such as tardive dyskinesia. The ethical and legal imperative of offering a less intrusive treatment was ignored. Benzodiazepines were not offered. Psychotherapy was not offered or mentioned in fifteen of the 30 cases. The providers claimed it does not work, even though that statement is blatantly false. The legal procedures can best be characterized as a sham where the patients are defenseless. The power imbalance and abuse were extreme and several of the psychiatrists who argued for forced treatment obtained court orders for administering drugs and dosages that were dangerous. Forced medication should be abandoned.

## Introduction

Involuntary civil commitment in the United States is a legal intervention. A judge or someone in a judicial capacity may order that a person with symptoms of a serious mental disorder who presents a danger to self or others can be confined to a psychiatric hospital or receive supervised outpatient treatment. Standards and procedures for commitment are provided by state law in every state. Involuntary commitment and involuntary medication proceedings must comport with due process protections under state and federal law.

In the State of Alaska, the Law Project for Psychiatric Rights (PsychRights) pursued a strategic litigation campaign against forced psychiatric drugging through its founder, James B. (Jim) Gottstein, Esq., and won two Alaska Supreme Court cases ruling Alaska’s forced drugging regime unconstitutional.

The first case was *Myers versus Alaska Psychiatric Institute* (1). In that case, the Alaska Supreme Court held that in order to be forced to take psychiatric medication against their will, the court had to find by clear and convincing evidence that (1) the patient does not have the capacity to give or withhold informed consent; (2) it is in the patient’s best interest to take medication, which means that the benefits outweigh the harms; and (3) there is no less intrusive alternative available.

The second case was *Bigley versus Alaska Psychiatric Institute* (2). The court held that forced drugging petitions must include the “Myers factors” which are (1) an explanation of the patient’s diagnosis and prognosis, and the symptoms with and without medication; (2) information about the proposed medication, it’s purpose and method of administration, possible harms (euphemistically called side effects) and benefits; (3) review of the patient’s history including medication history and previous harms; (4) an explanation of interactions with other drugs including over-the-counter drugs, street drugs and alcohol; and (5) information about alternative treatments and risks along with benefits.

On June 1, 2016, Peter C Gøtzsche testified in an involuntary medication proceeding under Alaska Statutes 47.30.839 held in Anchorage, Alaska, and in connection with that reviewed four AS 47.30.839 petitions. All four petitions were strikingly similar and failed to provide the information required in the *Bigley* case.

We therefore wished to investigate more formally if the legal predicates for the involuntary administration of psychotropic medication orders were uniformly lacking. We assembled two cohorts of 30 patients, one from Anchorage, Alaska and one from Copenhagen, Denmark.

This report describes our experience from Alaska. The results from the Danish cohort have been published (3,4).

## Methods and materials

We asked the court to provide its files for 30 consecutive AS 47.30.839 petitions from Anchorage with January 1, 2016 as the planned starting date. In Alaska, such files are normally confidential and to obtain them, Gottstein submitted a request that access be granted for our research while preserving confidentiality. It turned out to be very difficult to get access to such cases with objections from both the hospital and the Alaska Public Defender Agency, which represent almost all of the psychiatric patients against whom AS 47.30.839 petitions are filed. They objected to providing us with the files. It required two trips to the Alaska Supreme Court and over four years to be granted access to redacted files of these proceedings. Because of the delay, the start date for the files was changed to the most recent files as of the date access was granted.

When the cases took place, the commitment hearings were held mostly by Zoom due to the Covid-19 pandemic. In the court room or by Zoom, the judge, the prosecuting attorney representing the State of Alaska and Alaska Psychiatric Institute and a public defender representing the patient were present.

The patients themselves were not always present, but they had the choice of either being in the court room or attending the court hearing via Zoom.

The hearings had two parts. The first was whether the patient required mental health commitment. The second was whether a medication petition for involuntary drug administration should be granted.

Based on the written material available to us, we noted the judge’s ruling and evaluated if, based on the criteria from the *Myers* and *Bigley* cases, the petitions, hearings, and decisions complied with the following requirements:

1. Information was provided that documented that the patient could not provide informed consent;
2. The information about the psychiatric drugs the patient took or would be forced to take was accurate;
3. A less intrusive alternative was available;
4. The combination of drugs the patient took was safe;
5. The arguments for using force were reasonable and documented;
6. The patients’ rights were respected; and
7. There were striking similarities from case to case considering that the patients were different.

## Results

We were able to obtain access to 30 consecutive cases, which were heard in court between January 3, 2018 and August 19, 2020. We reviewed the cases to see whether or not there was compliance with the Alaska cases *Myers versus Alaska Psychiatric Institute* and *Bigley versus Alaska Psychiatric Institute*.

In all 30 cases the was a Notice of 30-day Commitment Hearing document which outlined the patient’s rights such as having representation by counsel, call experts, and the ability to appeal an involuntary commitment. In the following, we describe the results according to our seven requirements.

### 1 Information was provided that documented that the patient could not provide informed consent

We reviewed the cases as to whether the issue of informed consent was addressed at the time of the hearings for mental health commitment and medication orders. Under AS47.30.839(g), the requirement is that the patient does not have the capacity to give or withhold informed consent in order for the medication petition be granted. Informed consent is crucial because it is important to determine whether or not the patient is able to make an informed choice about making healthcare decisions. Consent also protects the patients against assault and battery in the form of unwanted medical interventions. Psychiatric medications have the potential for severe adverse effects. Informed consent is important for protecting the legal rights of the patients and also helps guide the ethical practice of medicine. A high standard of informed consent can safeguard the patients’ rights to autonomy and self-determination in respect to the individual.

For all the requirements to be fully met, there would have to be evidence that the patient did not have capacity to give or withhold informed consent. In 29 of the 30 cases, the commitment petition was granted. One case went to a jury trial and the jury found in favor of the patient.

AS 47.30.839(d) requires a Court Visitor be appointed to make recommendations to the court. The Court Visitor prepares a report and testifies in the hearings as to whether they believe the individual has capacity for informed consent in regard to taking medications.

In 27 cases (90%), the medication order was granted. For this to happen there would have had to be discussion about whether the individual could give or withhold informed consent. However, in 16 of the records (53%), there was no mention throughout the hearings regarding competency of the individual even though the judge sometimes made it clear that the hearing was held to determine the capacity of the individual and even though the Court Visitors were expected to mention this. In three of the cases, the patients were already deemed incompetent to stand trial and in one case the patient had a guardian.

In one case, there was an initial denial of the medication petition order because the judge was concerned about the testifying doctor. The court indicated that “the doctor was confused about whether or not the patient can give informed consent or not,” and the judge did not grant the medication petition citing the *Myers* case. However, the medication order was granted at a second hearing two weeks later because the judge felt confident in the Court Visitor who testified that the patient was not competent. The patient said he felt “knocked out” when he took medication and declined medications but there was testimony that the patient was improved with medications.

In another case, the patient herself testified at the hearing and said that she was competent to refuse medications. She preferred therapy, enjoyed the groups at the hospital but the judge granted the medication order. The psychiatrist testified that she was disorganized in her thinking, was not eating, and said she was unable to care for herself.

In a third case, the patient was deaf and blind, and his parents were guardians. The parents were no longer able to care for the patient and the only real option for placement was the Helen Keller Institute in New York. The judge was very concerned that the drugs would sedate the patient so much he would not be able to participate in the placement interview, thereby rendering the patient unable to be placed at the institute. The judge said, “I don’t have confidence in the doctor” and felt that the drugs were used in a way where the doctor would “give him this or that” in a capricious or arbitrary manner. Nonetheless the medication petition was granted.

Two patients expressed in court that they had a “right to refuse medication.” One patient agreed to take medication but only at lower doses. The Court Visitor testified that the patient had the ability to consent to treatment and was therefore competent to make the decision. The treating psychiatrist disagreed, and the medication petition was granted.

### 2 The information about the psychiatric drugs the patient took or would be forced to take was accurate

There was much discussion about which medications the psychiatrist intended to use, and in all cases, several drugs were proposed, usually a combination of a psychosis drug and a so-called mood stabilizer (which usually means lithium or an antiepileptic), sometimes adding a depression drug.

There was little concern about the harms. In only three cases, did the judge reject the polypharmacy (see below).

Typically, during the medication petition part of the hearing, the prescriber would describe the planned medications and their side effects, often mentioning the risk of a permanent neurologic disorder - tardive dyskinesia - and the rare possibility of neuroleptic malignant syndrome.

However, there was no contrary testimony to what the provider - a psychiatrist, physician assistant or nurse practitioner – recommended except for cross-examination by the public defender. While the public defenders all seemed to care about their clients, they had little to say to oppose the state attorney and they never presented an opposing psychiatrist or other expert witness. The only “proof” that the medication was in the person’s best interest was the unopposed opinion from the treating psychiatrist.

Thus, with a few exceptions, the hospital was able to direct their psychiatric practice in the manner they saw fit. There was no way to determine if what the providers argued regarding treatment was accurate, but some of the judges were experienced and used their own knowledge in some of the cases, e.g. questioning the need for so much medication and even at times the dose of medications. One judge thought that the patient was not psychotic, although the provider said so.

The patients’ experience with previous drug treatment was never taken into consideration even though it was extensive and although many expressed their opinions.

There was never any discussion about what types of drugs the patients preferred. The patients commented on the harms of the drugs, and one noted that she did not want lithium because it made her “shake physically” and she did not feel well on it.

One patient mentioned that Lamictal (lamotrigine) had helped him in the past, but the treating doctor did not want to use it. He said he would rather use an antipsychotic medication and the patient was started on Geodon (ziprazidone) 40 mg b.i.d., which is double the starting dose recommended by the FDA (5).

In another case, the treating psychiatrist said the patient did not have any side effects from the medication, but the court visitor testified that she had tardive dyskinesia. The psychiatrist became defensive and said, “She will experience neurological damage if she is not treated,” and the medication order was granted. The patient had involuntary mouth movements from years of Thorazine (chlorpromazine) treatment. The patient’s public defender argued that she suffered from a developmental disability and not a mental illness. It was suggested that she could get a reduced amount of medication due to her history of intellectual disability, but no concessions were made by her provider.

One patient was vehemently objecting to medication and noted that she had side effects that made her hands cramp. She requested a psychotherapist and said her son would pay for it. The provider argued that “There would be no therapeutic benefit from therapy.” The patient objected to medication because it “takes my feelings away,” but the provider said: “I will give you medications that get your feelings back,” which is blatantly wrong as all psychiatric drugs remove or dampen feelings. The patient said, “The medication will give me a heart attack and liver disease.” She had been on Depakote (valproate), and the FDA warns that the drug can cause fatal heart blocks and fatal hepatotoxicity (6). The provider requested two psychosis medications. In the medication petition hearing, the public defender quoted the *Myers* case, arguing that the patient should have the least intrusive treatment and should not be given two medications. The judge ordered that only one medication, Depakote should be given. The psychiatrist wanted to prescribe Depakote as he said it was the only medication that treated her bipolar disorder, but the judge order was that she would be treated with i.m. olanzapine if she refused to take Depakote and Ativan (lorazepam).

One patient experienced tremor from Haldol (haloperidol), a well-known harm of the drug, particularly at overdosage (7). The public defender requested limiting the dose to 30 mg orally daily. The provider, a nurse practitioner, requested up to 100 mg orally on a daily basis, which is an extreme dose. The recommended doses go up to 6 mg daily for moderate symptoms and 15 mg for severe symptoms. During that medication petition hearing, the prosecuting attorney for the Alaska Psychiatric Institute said that there was an instruction from the Alaska Supreme Court that courts should not micromanage petitions for medications. He also argued that 100 mg of Haldol was the medical standard of care, but the judge limited the dose to 30 mg.

In some instances, the medication amounts were duplicate; for instance, one individual was on oral Abilify (aripiprazole) 30 mg daily in addition to an injectable Abilify dose of 882 mg on a monthly basis. Generally, there is no need for oral and injectable medication of the same drug.

It was assumed that all the patients required medication. There was no discussion of the patients possibly doing better without medication or on reduced doses. All the treating psychiatrists said that the benefits outweighed the risks of the medications.

### 3 A less intrusive alternative was available

Consideration of alternative and less intrusive treatments along with the risks and benefits was a requirement from the *Myers* case. There weren’t really any alternatives offered while, in other areas of world, there are programs such as Open Dialogue in Europe and a facility called “Alternative to Meds” in Arizona. Benzodiazepines are much less toxic than antipsychotics but this option was not mentioned in any of the 30 cases.

In 15 cases, alternatives to drugs were mentioned such as psychotherapy or occupational therapy, but in every single case, the provider opined that it would not be helpful, even when the judge had asked if the patient could benefit from talk-based therapy.

The underlying assumptions were that all drugs are good and that all combinations of drugs are good. The dangers of the psychiatric medications were minimized and the plan was in all 30 cases to have the patients take medication, live in an assisted living facility or hospital without any thought of what could be done to improve their functional capacity and lives. For some, the heavy drugging regimen would render them incapable of getting employed or sustaining relationships.

### 4 The combination of drugs the patient took was safe

It was presumed that the medications only had positive effects and that side effects were uncommon. All the providers recommended at least two medications, sometimes three or four. There was no discussion about possible drug-drug interactions even though commonly prescribed drugs, e.g. proton pump inhibitors can reduce clearance of other drugs, which could result in overdoses of psychiatric drugs.

It was not considered if substance abuse contributed to the patients’ symptoms, although some of the patients had significant substance abuse histories and there was no discussion about possible drug-drug interactions with these substances.

### 5 The arguments for using force were reasonable and documented

The providers testified that emergency medication was needed at times and this was always requested, with no opposing testimony as to the requirements or documentation.

In 14 cases there was some reference to force being used. Several individuals were held down by the staff in order that they be injected. One psychiatrist testified that if a patient refused oral medications, he would be held down and injected. At times, the patients were threatened with injections but they acquiesced and agreed to the oral medication rather than receiving the injection.

### 6 The patients’ rights were respected

The patients’ rights were not respected and their thoughts, plans and wishes were never considered. One patient did not wish to take medications and the public defender made this clear in the closing argument in court and argued he may benefit from increased psychosocial support.

One patient did not want to take Haldol because it made him to groggy and “took away his feelings.” He told the doctor that Haldol calms him down in small doses and he agreed to low doses. The patient’s son requested Haldol not be used because it exacerbated his anger. The son noted that Haldol also caused lethargy and muscle cramps.

One patient stated that “they labelled me as schizophrenic but they can’t prove it.” She wanted to stay at the facility and prove to them that she did not need medication. She was probably going to be at the Alaska Psychiatric Institute for a year. The judge questioned whether or not she was psychotic. She herself testified and explained her side effects. Two antipsychotics were requested but only one was granted.

One patient wanted to see how she did without medications and prove to the providers that she could do well without medications. The possibility of a trial without medication that this patient requested was not considered.

Some of the patients had reduced cognition secondary to the mental health disorder or medication. Eight patients said that they did not need medication and eleven other patients objected to the medication due to its harms. One said, “I’ve been drugged out of my mind.” One patient declined medication because she believed she was a psychiatrist. All patients had a medication order granted by the court.

### 7 There were striking similarities from case to case considering that the patients were different

All the patients were presumed to have a mental health disorder. There was no consideration as to the possibility that the person’s symptoms were secondary to their history of substance abuse or other condition. Many of them seemed to be experiencing withdrawal symptoms after they had stopped medication, sometimes abruptly, with subsequent development of psychosis, but these symptoms were always thought to be part of their primary psychiatric disorder. There was no documentation that any of the patients were warned about the possibility of severe withdrawal symptoms if the psychiatric medications were abruptly discontinued.

## Discussion

The patients’ human rights were systematically violated and the precedents stemming from the *Myers* and *Bigley* Supreme Court cases in Alaska were consistently ignored.

The psychiatrists got away with the argument that, in their opinion, it was in the patients’ best interests to be forcefully treated with a psychosis drug. This argument is invalid, and a healthcare professional cannot be excused for not knowing about the science or for ignoring it. Psychosis drugs do not have any specific effects against psychosis and it is therefore misleading to call them antipsychotics. They work the same way in patients, human volunteers, and animals, basically by knocking people down (8) so that they cannot function, which is why their original name, major tranquilizers, was more appropriate.

It is well-known that placebo-controlled trials of psychosis drugs are highly flawed. One of the reasons is that patients recruited for the trials were already in treatment with such a drug before randomization (9,p.44). Psychosis pills can cause psychosis, known as supersensitivity psychosis or oppositional tolerance, even during continued treatment (8p.45,10). The drugs decrease dopamine levels, and the number of dopamine receptors goes up to compensate for this. If the drugs are suddenly stopped, the response can very well be a withdrawal psychosis. The trials therefore only show what happens when patients randomized to placebo get harmed by a cold turkey.

Another important bias is the lack of effective blinding because of the drugs’ conspicuous harms. When atropine is added to the placebo to mimic some of the harms of depression pills, the effect is markedly smaller than in usual placebo-controlled trials (11).

Virtually all of these trials are carried out by the drug industry, and a third important bias is serious manipulation with the data analysis or outright fraud (9,12).

Despite these formidable biases, the effect reported in the placebo-controlled trials of recent drugs submitted to the FDA was only 6 points on the Positive and Negative Syndrome Scale (13), whereas the minimally clinically relevant effect corresponds to about 15 points on this scale (14).

The huge CATIE trial, financed by the US National Institute of Mental Health is also telling of the poor effect of the drugs (15). It randomised 1493 “real world” patients with schizophrenia to olanzapine, quetiapine, risperidone, or ziprasidone, or to an old drug, perphenazine, marketed in 1957. The primary outcome was a very reasonable one, time to discontinuation for any reason, which reflects both the benefits and the harms of the drugs. After 18 months, only 26% of the patients were still on the randomized drug, and perphenazine was not worse than the newer drugs and did not produce more extrapyramidal harms than these agents, even though this is usually claimed (9).

The final blow to the argument that it is in the patients’ best interest to be treated with psychosis drugs is that both randomized trials with long-term follow-up and carefully conducted observational studies comparing treated with untreated patients have shown that more patients get rehospitalized and end up on disability pension when they receive psychosis drugs (9,16-18) (this research is summarised in ref. 9).

In addition to ignoring the *Myers* requirements, the court violated the principles laid down in the United Nations Convention on the Rights of Persons with Disabilities (19). The Convention has specified that member states must immediately begin taking steps towards the realization of the patients’ human rights by developing laws and policies to replace regimes of substitute decision-making by supported decision-making, which respects the person’s autonomy, will, and preferences (19). The convention has been ratified by virtually all countries except the United States, but this cannot be an excuse for not living up to it. We have an obvious ethical obligation to respect the patients and involve them in our decisions, and this ethical imperative cannot be suspended. Being psychotic does not mean that the patients are incapacitated as regards their views on and experiences from being treated with psychiatric drugs (19).

Polypharmacy of patients with psychosis was very common but it increases their risk of dying markedly. It is particularly bad medicine to try to force two psychosis drugs on a patient, as psychosis drugs double the risk of dying (9p.47,20) and as this harm is clearly dose related (21-25). Antiepileptics also increase the risk of dying, e.g. they double the risk of suicide (26).

There are no randomized trials that show that polypharmacy leads to better outcomes than treatment with just one drug or with psychotherapy. In addition, very little is known about interactions between the medications. Polypharmacy can be considered an off label or experimental treatment that should not be used, particularly not involuntarily.

The argument that the patients’ brains will be damaged if they are not treated with psychosis drugs, which was used in the court, is also commonly seen in textbooks (9). but it is totally wrong. Psychosis pills can cause irreversible brain damage (9,27,28), and it has never been shown that the psychosis per se can cause brain damage. One type of brain damage is tardive dyskinesia, which psychiatrists very often ignore. Among 58 consecutively admitted patients with acute psychosis, 48 of whom were treated for at least one week with psychosis drugs, the researchers found 10 patients with tardive dyskinesia, but the psychiatrists only made this diagnosis in one of them (29). It took psychiatry 20 years to recognise tardive dyskinesia as an iatrogenic illness (30), even though it is one of the worst harms of psychosis drugs and affects about 4-5% of patients every year (31), which means that most patients in long-term treatment will develop it.

The internationally established principle, confirmed also in the *Myers* case, of offering a less intrusive treatment was totally ignored. Benzodiazepines are far less dangerous than psychosis pills and even seem to work better for acutely disturbed patients (32), but they were never offered.

Psychotherapy was not offered either and in all 15 cases where this issue was raised, the providers claimed it does not work. This is totally wrong (9,33-38). A systematic review of seven trials showed that cognitive behavioral therapy can reduce the risk of developing psychosis by 50% (34), which is a huge effect.

It was not until 2014 that the first trial of psychotherapy in people with schizophrenia who were not on psychosis drugs was published (36). All the patients had declined to be treated with such drugs. The effect size was 0.46 compared to treatment as usual, about the same as that seen in seriously flawed trials comparing psychosis pills with placebo, which is a median of 0.44 (39). These and other results, e.g. those obtained with the Open Dialogue approach in Lappland (9,40). compared to treatment as usual (41), means that psychotherapy is far better than pills, particularly in the long run, as psychotherapy can help the patients live more normal lives while psychosis pills do the opposite (9,17,18,40,41).

Psychotherapy for schizophrenia even seems to be cost-effective. According to a NICE guideline from 2012, a systematic review of the economic evidence showed that cognitive behavioral therapy improved clinical outcomes at no additional cost, and economic modelling suggested that it might result in cost savings because of fewer hospital admissions (37,38).

### Comparison between Alaska and Denmark

There were striking similarities between the human rights violations in Alaska and Denmark. Gøtzsche’s review of 30 consecutive cases from the Danish Psychiatric Appeals Board showed that not in a single case was clear and convincing evidence presented that the proposed treatment was in the patient’s best interest (3,4).

According to Danish law, forced medication should be with drugs with the fewest possible adverse effects, but this condition was violated in 29 cases (97%). In 7 cases (23%), where the Appeals Board disagreed with an earlier decision made by the Psychiatric Patients’ Complaint Board and resolved that the conditions for forced treatment with a psychosis drug had not been met, the issues were formal and minor, and the Appeals Board argued nonetheless that force was justified because the patient was insane and that the prospect of cure or a significant and decisive improvement in the condition would otherwise be significantly impaired.

As noted above, both arguments are invalid. Like in Alaska, the Appeals Board seemed mainly to have a cosmetic function, rubber stamping what the psychiatrists’ wanted and focusing on uncontroversial issues it could easily check and not on what was best for the patients. In both countries, the outcome was a foregone conclusion, and the patients’ desires, fears, wishes, and experiences were totally ignored. Supported decision making was never an issue and there were no plans to improve the patients’ ability to function and to help them lead a full and purposeful life.

The patients’ reactions were also very similar. Several patients expressed fear of dying because of the forced treatment. These very valid concerns were ignored or cited as proof the person was delusional, even though one patient said: “my father died because of intoxication with psychiatric drugs” (4). Some patients have seen fellow patients suddenly drop dead because of the psychosis pills forced upon them, and some have even died themselves shortly afterwards (42).

Several patients, both in Alaska and Denmark had clear signs of tardive dyskinesia, which were discounted by the psychiatrists who ascribed the side effects to their illness even though schizophrenia cannot cause tardive dyskinesia. The psychiatrists recommended continued treatment with psychosis drugs despite the serious harms they had caused.

Neither in Alaska nor in Denmark was the issue of withdrawal symptoms ever brought up even though some of the patients in both countries suffered from them, which seemed to have led to lack of control, aggression and sometimes to a withdrawal psychosis. The psychiatrists never considered that the patients’ symptoms were due to drug withdrawal.

Akathisia was also ignored even though this drug harm is dangerous, as it increases the risk of suicide and violence, including homicide (9,12). An expert confirmed our suspicion that a patient had developed akathisia on aripiprazole; but on the same page this expert – a high-ranking member of the board of the Danish Psychiatric Association – recommended forced treatment with this drug even though it had been stopped because of the akathisia (4). This is serious medical malpractice.

The patients or their disease were blamed for virtually everything untoward that happened. We did not see a single admission that it was the psychiatrist or other staff who had escalated a situation by their insistence that the patients be treated with drugs they could not tolerate or did not want or with other forced measures. We found several clear examples that it was the impending use of force that made the patients aggressive (4). In five of the Danish cases, the explicit purpose of forced treatment was not to benefit the patients but to prevent them from disturbing the staff and other patients.

In Denmark, we had reservations about the psychiatrists’ diagnoses of delusions in nine cases (4). For example, when a patient rejected olanzapine totally, this was called a persecutory delusion; another patient who became “hotheaded and difficult to communicate with” as soon as an antipsychotic was mentioned, was called “paranoid and conspiratorial about how we rally against him”. One patient with clear signs of tardive dyskinesia was said to have psychotic misconceptions about the “postulated side effects;” when a patient mentioned that she was served meat even though she was a vegetarian, this was interpreted as a delusion; and a patient who wanted to complain about being subjected to forced medication was also called delusional.

The disconnect between the views of the psychiatrists and their patients was vast. In all 21 cases in Denmark where there was information about the effects of previous drugs, the psychiatrists stated that psychosis drugs had had a good effect whereas *none* of the patients shared this view (4). Seven patients asked for a psychologist, but this seemed not to have been granted. Even when the patients had explained that they could not tolerate the drugs, they were forced upon them to “preserve their health.” One patient noted that her medication caused psychosis, but to treat the psychosis, the dose kept getting increased and she became overmedicated and unable to manage her life. Another patient noted that one definition of madness is administering poison and expecting your victim to heal.

In Alaska, one patient felt “near death” while taking drugs but her psychiatrist said, “She is improved on medications.” She reported constipation, jerking muscles and inability to urinate and declined medications: “Let me stay here long enough to prove that I don’t need medication.” Even the judge questioned if she was psychotic. She testified herself, but the judge did not respect her wishes but allowed treatment with one psychosis drug, olanzapine, one of the worst psychosis drugs (9), instead of the requested two drugs, the other being quetiapine.

The approach in both countries was to focus on heavy medication instead of recovery, which meant that many patients would need to live permanently under assisted housing conditions with no real future. A Danish patient who had become lethargic while receiving three psychosis drugs simultaneously would rather go to jail than be given drugs. And an Alaskan patient said, “I have more rights in jail than here” (in the Alaska Psychiatric Institute) and that, “You can get in and out faster when you are in jail.”

The legal procedures in both countries can best be characterized as a sham where the patients are defenseless. The power imbalance and abuse we found were extreme and several of the psychiatrists who argued for forced treatment obtained court orders for administering drugs and dosages that were dangerous. In the US, it has been documented that psychiatrists, with the full understanding and tacit permission of trial judges, regularly lie in court to obtain involuntary commitment and forced medication orders (12,43). Mendacious information is routinely included in petitions and testimony also in Alaska. Gottstein’s book, *The Zyprexa Papers*, details this with some particularity regarding the forced drugging proceedings against Bill Bigley (44).

In Norway, the Ombudsman concluded in December 2018 that the Psychiatry Act had been violated in a specific case because the randomized trials showed that the probability of achieving the intended improvement was low (45).

## Conclusions

The systematic violation of the rights of psychiatric patients and the discrimination against them is a global problem and the harms inflicted on the patients is immense. Forced medication must be abandoned.

## Data Availability

The anonymised raw data can be obtained from the authors.

## Acknowledgment

We thank Jim Gottstein for his invaluable help with getting access to documents.

## Contribution of each author

PCG wrote the protocol for the study; GT interpreted the raw data; and both authors contributed to writing the manuscript.

## Data sharing

The anonymised raw data can be obtained from the authors.

